# Evaluation of the Abbott Panbio™ COVID-19 antigen detection rapid diagnostic test among healthcare workers in elderly care

**DOI:** 10.1101/2022.10.04.22280700

**Authors:** Andrea Eikelenboom-Boskamp, Martijn den Ouden, Theun de Groot, Tim Stobernack, Heiman Wertheim, Andreas Voss

**Author notes:** Corresponding Author Andrea Eikelenboom-Boskamp, (AEB).

## Abstract

**Background:** Coronavirus disease 2019 (COVID-19) has been especially dangerous for elderly people. To reduce the risk of transmission from healthcare workers to elderly people, it is of utmost importance to detect possible severe acute respiratory syndrome coronavirus 2 (SARS-CoV-2) positive healthcare workers as early as possible. We aimed to determine whether the Abbott Panbio™ COVID-19 antigen detection rapid diagnostic test (Ag-RDT) could be used as an alternative to reverse transcription-quantitative polymerase chain reaction (RT-qPCR). The second aim was to compare the cycle threshold (Ct) in RT-qPCR with the results of the Ag-RDT.

**Methods:** A prospective diagnostic evaluation of the Abbott Panbio™ COVID-19 Ag-RDT among healthcare workers across three elderly care facilities as well as home-based elderly care workers who met clinical criteria for COVID-19 during the second wave of the COVID-19 pandemic. Per healthcare worker, the first nasopharyngeal swab was obtained to perform the Ag-RDT and the second swab for RT-qPCR. A Ct-value of < 40 was interpreted as positive, ≥ 40 as negative.

**Results:** A total of 683 healthcare workers with COVID-19 symptoms were sampled for detection of SARS-CoV-2 by both Ag-RDT and RT-qPCR. Sixty-three healthcare workers (9.2%) tested positive for SARS-CoV-2 by RT-qPCR. The overall sensitivity of Ag-RDT was 81.0% sensitivity (95%CI: 69.6-88.8%) and 100% specificity (95%CI: 99.4-100%). Using a cut-off Ct-value of 32, the sensitivity increased to 92.7% (95% CI: 82.7-97.1%). Negative Ag-RDT results were moderately associated with higher Ct-values (*r* = 0.62) compared to positive Ag-RDT results.

**Conclusion:** The Panbio™ COVID-19 Ag-RDT can be used to quickly detect positive SARS-CoV-2 healthcare workers. Negative Ag-RDT should be confirmed by RT-qPCR. In case of severe understaffing and with careful consideration, fully vaccinated healthcare workers with Ag-RDT negative results could work with a mask pending PCR results.

## INTRODUCTION

Elderly care facilities are high-risk settings for transmission of COVID-19 to and among residents and healthcare workers. Residents are at a higher risk of developing severe infection due to age and comorbidities. Early detection of SARS-CoV-2 among healthcare workers and vulnerable residents and rapid contact tracing are critical to reduce the risk of SARS-CoV2 transmission and COVID-19-associated morbidity and mortality.(1,2)

Given the epidemiology of the COVID-19 epidemic and changes in guidelines for elderly-care facilities (3), there is a strongly increasing test demand for SARS-CoV-2. Therefore, antigen detection rapid diagnostic tests (Ag-RDTs) with fast results may be an inexpensive, scalable solution. The ministry of Health, Welfare and Sport (VWS) and National Institute for Public Health and the Environment (RIVM) have selected five Ag-RDTs for clinical validation based on the technical validation and potential availability (4). The Panbio™ COVID-19 Ag rapid test is one of the selected Ag-RDTs and generates a result within 15 minutes. This Ag-RDT was ordered by the elderly care facilities when it became available within the regular ordering system. The manufacturer of the Panbio™ COVID-19 Ag rapid test reported 93.3% sensitivity and 99.4% specificity in asymptomatic people with high viral load. The diagnostic performance of this test among healthcare workers is not known.

Our objective was to determine whether the Panbio™ COVID-19 Ag-RDT can be used as an alternative to reverse transcription-quantitative polymerase chain reaction (RT-qPCR) among healthcare workers working in elderly care. Moreover, the cycle threshold (Ct) in RT-qPCR, which is needed to detect virus and inversely proportional to the viral load, was determined.

## METHODS

### Study design and population

Between November 2020 and January 2021, we conducted a prospective diagnostic evaluation of the Panbio™ COVID-19 Ag rapid test determined against RT-qPCR, which is considered as the ‘golden standard’, among healthcare workers working in elderly care facilities as well as in home-based elderly care who met clinical criteria for COVID-19 (5). Written information about this study was provided by email to all healthcare workers of the organization and verbal information was given on the spot when healthcare workers came to test. The reasons of healthcare workers who did not want to participate were not registered given the extra workload.

### Procedures

#### Training of personnel

Prior to the start of the study, personnel of the elderly care organization were trained to perform the Ag-RDT in accordance with the manufacturer’s protocol.

#### Nasopharyngeal swabs

Per healthcare worker, two nasopharyngeal swabs were obtained consecutively by dedicated personnel of the organizations themselves wearing personal protective equipment. The first nasopharyngeal swab was taken to perform the Ag-RDT and the second nasopharyngeal swab for RT-qPCR by the laboratory.

#### Panbio TM COVID-19 Ag rapid test

Panbio™ COVID-19 Ag rapid test device by Abbott (Lake Country, IL, U.S.A) is a membrane-based immunochromatography assay which detects the nucleocapsid protein of SARS-CoV-2 in nasopharyngeal samples. Collected swabs were transferred into dedicated sample collection tubes containing a lysis buffer provided with the test kit. Samples were processed on site, directly after collection. After 15 minutes of assay initiation, tests were interpreted. The test results were documented on the questionnaire as well as the image of the result window on the test device for processing and analysing the data by the researcher. The laboratory analysts involved in doing RT-qPCR were not informed about the result of the Ag-RDTs.

#### RT-qPCR

Nasopharyngeal swabs were collected using ∑-Transwab® in 1 ml liquid Amies medium and PCR was conducted in a certified clinical laboratory with procedures validated in accordance with the NEN-EN-ISO 15189 standard. Nucleic acid (NA) extraction was performed using MagNA Pure 96 DNA and Viral NA Small Volume Kit and MagNA Pure 96 Instrument (Roche Diagnostics GmbH, Mannheim, Germany) by following the manufacturer’s instructions. Before NA extraction, the internal control phocine distemper virus (PhDV) was added to the sample via Xiril robotic workstations (Roche), while another Xiril workstation was used for PCR setup by pipetting 10 µl of NA with 10 µl of master mix containing 5 µl TaqMan® Fast Virus 1 Master Mix (Thermofisher Scientific) and 5 µl of primers and probes (Supplementary Table 1), targeting SARS-CoV-2 E-gene and PhDV. Thermal cycling was performed in a LC480-II instrument (Roche) with 1 cycle of reverse transcription at 50°C for 5 min followed by 1 cycle of PCR activation at 95°C for 20 sec, followed by 45 cycles of 95°C for 3 sec and 60°C for 30 sec. Data analysis was performed using Roche FLOW software (Roche) and a Ct-value of < 40 was used to interpret results as positive.

#### Survey

Healthcare workers were asked to fill out a questionnaire regarding (onset of) symptoms, risk of exposure to SARS-CoV-2 and history of SARS-CoV-2 positivity. Dedicated personnel who performed the Ag-RDT completed the questionnaire with the result of the Ag-RDT.

#### Scenarios for test results

In addition to current local guidelines for prevention of COVID-19, scenarios were described for testing either positive or negative Ag-RDT as illustrated in Figure 1.

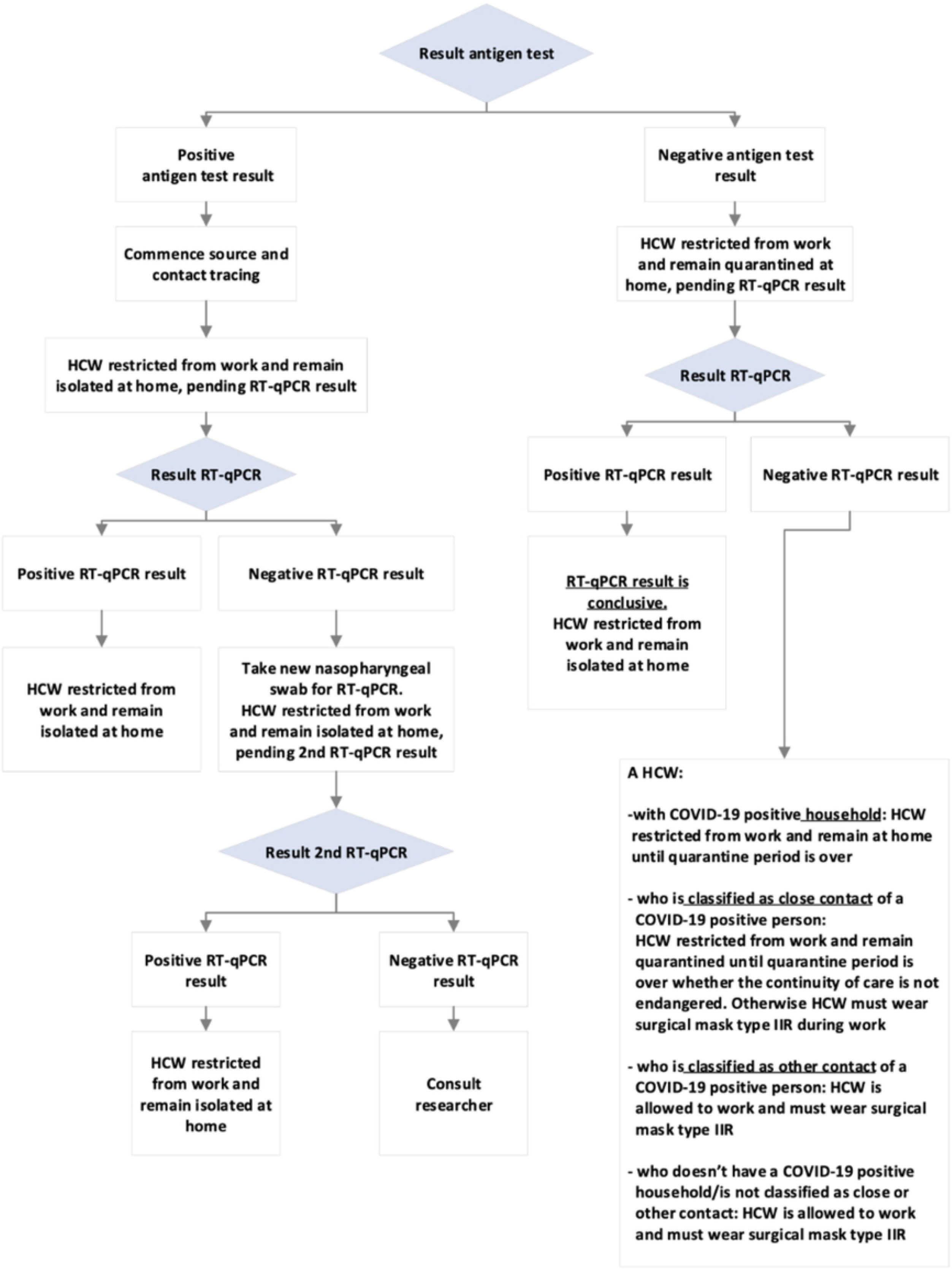

### Statistical analysis

Categorical variables were expressed as counts and percentages. Continuous variables were expressed as mean, median, interquartile range (IQR) and/or minimum-maximum (min - max). Difference testing for comparisons of groups was performed with Fisher’s exact tests for categorical variables and Mann-Whitney U tests for non-normally distributed continuous variables. Two-sided p-values less than or equal to 0.05 were considered statistically significant for all variables, with the exception of duration of symptoms and Ct-values for which a one-sided test was applied. Risk ratios were calculated to determine effect sizes of symptoms for positive SARS-CoV-2 results by RT-qPCR.

A Ct-value < 32 for E-gene, which is associated with culturable virus in nasopharyngeal specimen and therefore considered as infectious (6), was applied to the analysis mentioned below.

Association (*r*) between Ag-RDT results and Ct-values was calculated from the z-score of the Mann-Whitney U test. Association between COVID-19 symptoms and Ct-values was determined using Spearman’s Rho (ρ). Sensitivity, specificity and predicted values of the Ag-RDT were calculated with 95% confidence intervals (CI) using the RT-qPCR as the ‘gold standard’. The level of agreement between the tests was evaluated using Cohen’s kappa score.

Analyses were performed with SPSS statistics 27 (IBM), whereby 95% CI were calculated using OpenEpi version 3.0.3 (http://www.openepi.com/).

### Ethical approval

The study was reviewed (File number CMO: 2020-7083) by the ethics committee of the Radboud University Medical Centre, which decided that the study is not subject to the Medical Research Involving Human Subjects Act and did not require full review by an accredited Medical Research Ethics Committee. All participants have provided written informed consent.

## RESULTS

### Healthcare workers’ characteristics

A total of 683 healthcare workers with COVID-19 symptoms were sampled for detection of SARS-CoV-2 by both Ag-RDT and RT-qPCR. Based on RT-qPCR, 63 healthcare workers (9.2%) tested positive for SARS-CoV-2 from 11 November 2020 to 15 January 2021. The mean age of the respondents was 43.2 years (median 46.0; IQR 24.0, min – max: 16-65 years) of which 641 (93.9%) were female.

Six (9.5%) of the 63 SARS-CoV-2 positive healthcare workers reported to have tested SARS-CoV-2 positive previously. Results of the samples from these six healthcare workers were included in the diagnostic performance of the Ag-RDT. The reported symptoms were excluded from analyses.

Compared to negative healthcare workers, SARS-CoV-2 positive healthcare workers reported the following significantly more often: fever, flu-like symptoms (headache, muscle pain and/or fatigue), loss of taste or smell, exposure to SARS-CoV-2 positive household and close contact with SARS-CoV-2 positive person. The mean number of days between symptom(s) onset and tests among SARS-CoV-2 positive and negative healthcare workers by RT-qPCR were 2.0 and 2.2 days, respectively. Median days were equal in both groups, namely 1.0 days. These data are shown in Table 1.

Among SARS-CoV-2 positive healthcare workers, there was a statistically significant association between the presence of fever and Ct values < 32 (p = 0.0044); however, the association was consi

**Table 1.** Healthcare workers characteristics

|  | Total<br>N (%) | SARS-CoV-2<br>positive by<br>RT-qPCR, N (%) | SARS-CoV-2<br>negative by<br>RT-qPCR<sup>1</sup>, N (%) | Risk ratio for<br>cohort positive<br>by RT-qPCR<br>(95%CI) | P value<sup>a</sup> |
| --- | --- | --- | --- | --- | --- |
| **COVID-19 symptoms** | 683 |  |  |  |  |
| Fever | 94 (13.8) | 21 (22.3) | 73 (77.7) | 3.1 (1.9-5.0) | <0.001 |
| Shortness of breath | 70 (10.3) | 9 (12.9) | 61 (87.1) | 1.5 (0.8-2.8) | 0.276 |
| Respiratory symptoms | 530 (77.8) | 43 (8.1) | 487 (91.9) | 0.6 (0.4-1.0) | 0.078 |
| Headache, muscle pain, fatigue | 376 (55.2) | 47 (12.5) | 329 (87.5) | 2.4 (1.4-4.1) | 0.001 |
| Loss of taste or smell | 45 (6.6) | 11 (24.4) | 34 (75.6) | 3.0 (1.7-5.3) | 0.001 |
| Diarrhea | 61 (9.0) | 3 (4.9) | 58 (95.1) | 0.5 (0.2-1.6) | 0.351 |
| Hayfever | 36 (5.3) | 3 (8.3) | 33 (91.7) | 0.9 (0.3-2.7) | 1.000 |
| Unknown | 2 (0.3) | 0 | 2 (100) | N/A | N/A |
| **Exposed to SARS-CoV-2**<br>**positive person** |  |  |  |  |  |
| Household | 27 (4.0) | 9 (33.3) | 18 (66.7) | 4.0 (2.2-7.3) | <0.001 |
| Close contact<sup>b</sup> | 132 (19.3) | 20 (15.2) | 112 (84.8) | 1.9 (1.2-3.2) | 0.018 |
| Other contact<sup>c</sup> | 63 (9.2) | 8 (12.7) | 55 (87.3) | 1.4 (0.7-2.8) | 0.359 |
| Other reason for quarantine | 48 (7.0) | 1 (2.1) | 47 (97.9) | 0.2 (0.03-1.5) | 0.115 |
| Mean days between symptoms<br>onset and both tests [median,<br>SD] | 2.2 [1.0 (2.6)] | 2.0 [1.0 (2.3)] | 2.2 [1.0 (2.6)] | N/A | N/A |

a. Fisher's exact test, two-sided
b. Close contact: any individual within 1,5m distance of an infected person for at least 15 minutes or high-risk exposure less than 15 minutes
c. Other contact: any individual at a distance of more than 1,5m of an infected person in the same room for at least 15 minutes

dered as weak (ρ = 0.27). For other reported symptoms, no significant differences in Ct-values were identified. Data not shown.

### Diagnostic performance of the Ag-RDT

As presented in Table 2, 51 healthcare workers tested positive both by RT-qPCR as well as by Ag-RDT. Twelve healthcare workers had negative Ag-RDT results but positive RT-qPCR results, leading to an overall sensitivity of 81.0% (95%CI: 69.6-88.8%). When using a cut-off Ct-value of 32 instead of 40, the sensitivity of the Ag-RDT increased to 92.7% (95% CI: 82.7-97.1%). False-positive Ag-RDT results were not found, resulting in a specificity of 100% (95%CI: 99.4-100%).

**Table 2.**
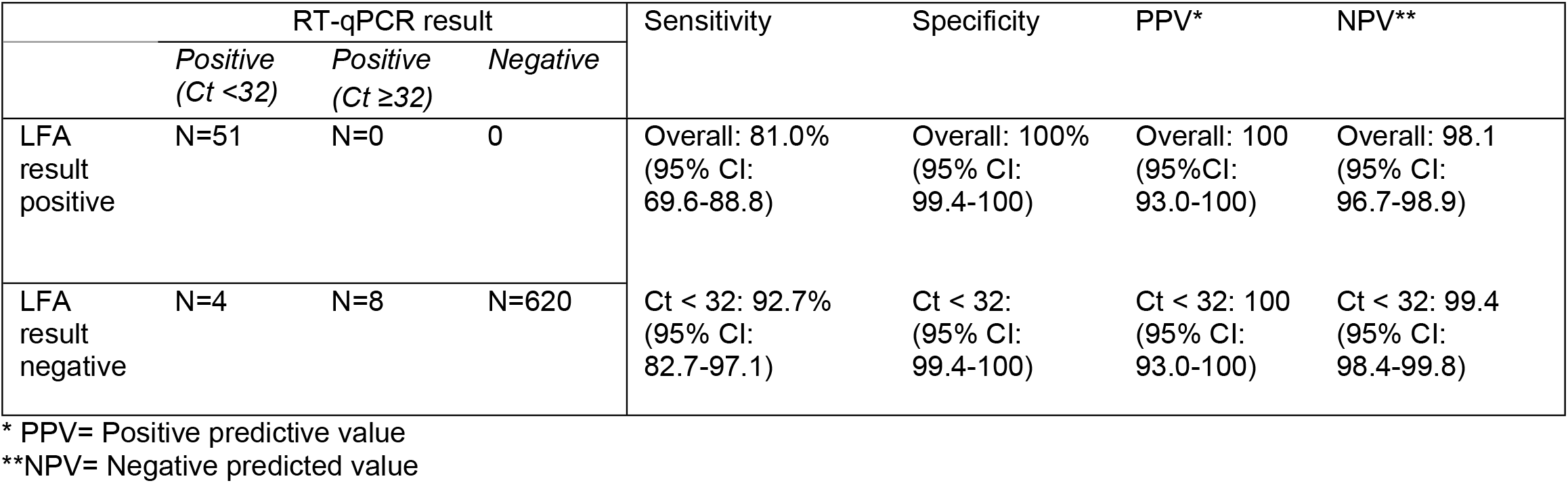
Diagnostic performance of Panbio™ COVID-19 Ag-RDT

As illustrated in Figure 3, the median Ct-value was significantly lower among the group of 51 healthcare workers with both positive Ag-RDT and PCR results compared to the group of 12 healthcare workers with negative Ag-RDT and positive PCR results, 20.61 and 32.34 respectively (p < 0.001). Negative Ag-RDT results were moderately associated with higher Ct-values (*r* = 0.62) compared to positive Ag-RDT results. The minimum and maximum Ct-value among the group with negative Ag-RDT and positive PCR results were 23.73 and 36.00, respectively. The particular Ct-values from the four healthcare workers with negative Ag-RDT and positive PCR result with Ct-value < 32 were 23.73, 24.11, 27.08 and 30.64. Among healthcare workers with both positive Ag-RDT and PCR results, minimum and maximum values were 15.00 and 29.75, respectively.

The positive predicted value (PPV) and negative predicted value (NPV) in this study cohort with a prevalence of 9.2% were 100% (95%CI: 93.0-100) and 98.1% (95%CI: 96.7-98.9), respectively. Using a cut-off Ct-value of 32, the PPV and NPV were 100% (95%CI: 93.0-100) and 99.4% (95%CI: 98.4-99.8), respectively. The overall negative predicted values for different prevalence are shown in Figure 2. An almost perfect agreement (Cohen’s kappa score = 0.885) was found between the two tests (p < 0.001), and when working with a cut-off Ct-value of 32 the Cohen’s kappa increased to 0.959.

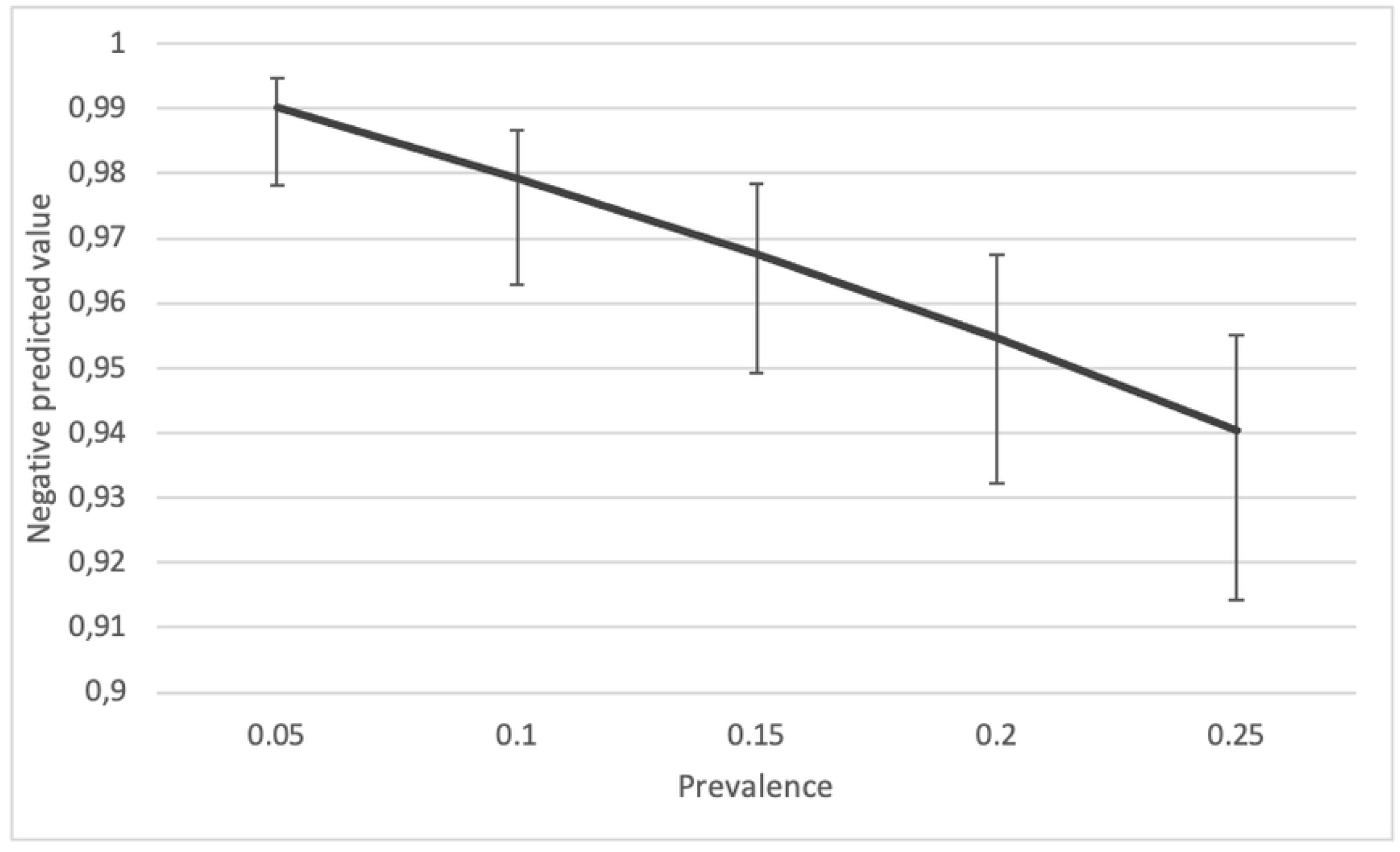

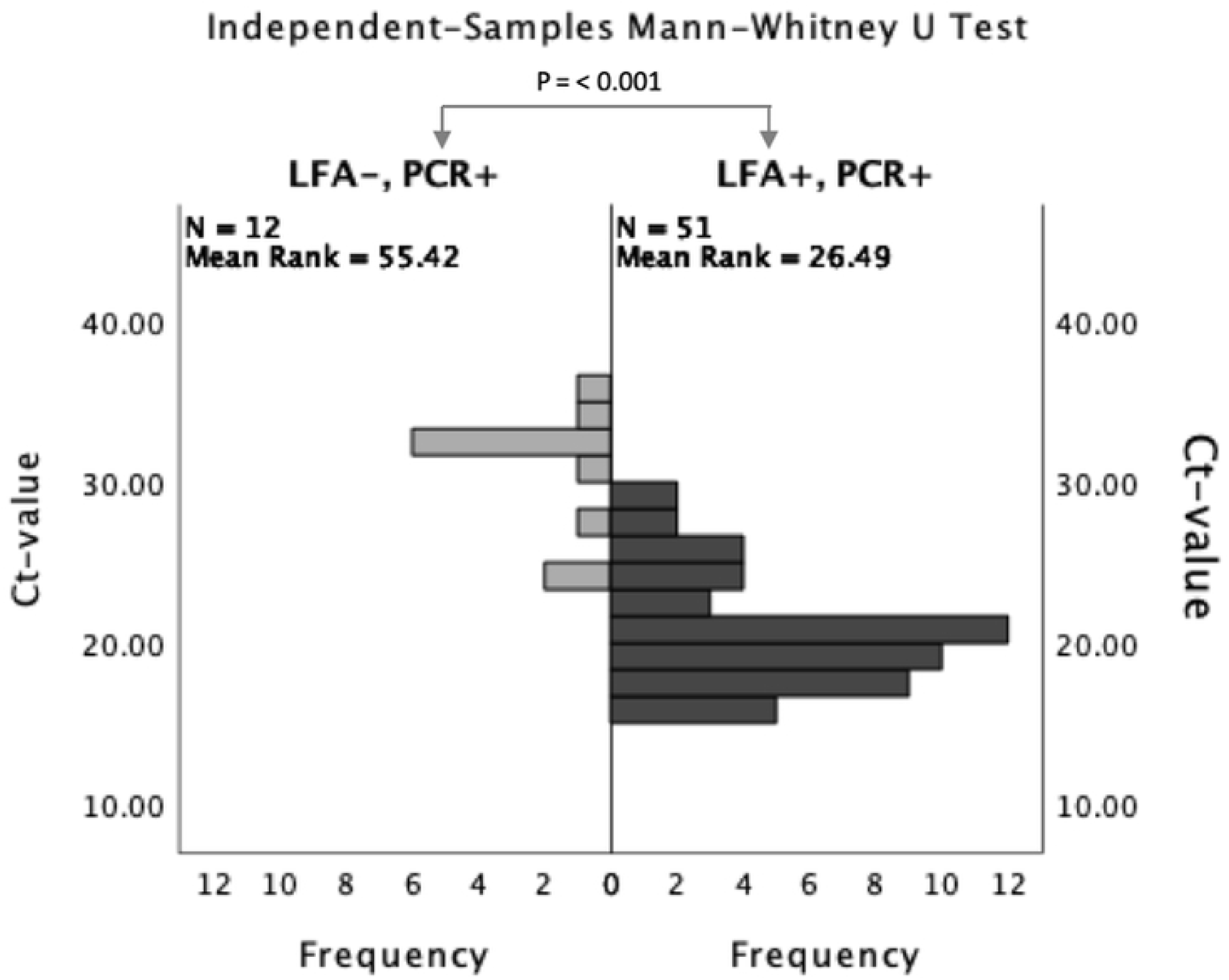

## DISCUSSION

Our findings have shown that the Panbio™ COVID-19 Ag rapid test can be considered a useful Ag-RDT among healthcare workers. Positive results become quickly available and do not need to be confirmed by RT-qPCR. Contact tracing could be started appropriately and immediately. Negative Ag-RDT results should be confirmed by RT-qPCR even with a reduced prevalence of SARS-CoV-2 in this setting where healthcare workers take care of residents who are at risk of developing severe infection.

In our study, the overall agreement between RT-qPCR and the Ag-RDT was almost perfect (κ = 0.885) and increased when using a cut-off Ct-value of 32 (κ = 0.959). Positive Ag-RDT results were found in 81% of positive RT-qPCRs. In 19% of cases, false-negative results were found; however, by considering Ct-values of ≥ 32 as not infectious, false-negative results were decreased to less than 8%.

Several studies have evaluated Panbio Ag-RDT as a useful test in different settings using different cut-off Ct-values as a measure for infectiousness (7–10). In our laboratory, primer sets targeting SARS-CoV-2 E-gene were used. Hence, we applied a cut-off Ct-value of 32 as measure for infectiousness based on a study from Huang et al. (6) to gain insight into the effect of using this cut-off value to the diagnostic performance of our Ag-RDT and to determine associations with COVID-19 symptoms. Unfortunately, SARS-CoV-2 positive healthcare workers were still prohibited from working because an (inter)national consensus on cut-off Ct-values has not yet been defined. In view of the results of our study, the probability of being infectious when having a negative Ag-RDT result is still low. Therefore, in cases of severe understaffing and pending the PCR results, it may be appropriate to allow fully vaccinated healthcare workers with a negative Ag-RDT, to work as long as they wear a face mask for the entire shift (including during breaks separate from colleagues). Given the vulnerability of the patients in this setting, these policies should be carefully considered. In addition, new SARS-CoV-2 variants may be more contagious than previous variants and vaccination may be less effective against new variants. For example, the delta variant seemed to be more than twice as contagious as previous variants (11), and vaccination was less effective at preventing transmission of the delta variant by vaccinated people than it was with the alpha variant. (12)

An important drawback of this Ag-RDT is that in case of a negative result a second nasopharyngeal swab is needed for RT-qPCR. In our study, all participants provided written consent; however, we do not know if the healthcare workers who did not provide written consent refused to participate in the study because it involved the double-swab method. It does not seem to be deterrent, given the number of participants in the study period. We think that the advantage of obtaining a positive SARS-CoV-2 result rapidly could outweigh the inconvenience and discomfort using a second swab. Despite the user-friendliness of the Ag-RDT, we recommend that this test be performed by trained and dedicated personnel to achieve a high level of accuracy. high performance.

An important point to note is that our study was carried out in the common-cold/flu(-like) season, during which many healthcare workers have respiratory symptoms, as well as in a period with a high prevalence of SARS-CoV-2. In such a situation and also in a local outbreak setting, it could be beneficial, in close cooperation with a medical microbiology lab, to establish or maintain a test lane or a local test-team that uses Ag-RDTs. An additional advantage is that results can be easily tracked in order to keep an overview of SARS-CoV-2 infections on an organization-wide level. However, to determine whether this test policy could be beneficial, it is important to take into account the turnaround time for SARS-CoV-2 results by PCR when testing at the Municipal Public Health Services (in Dutch: GGD).

## Data Availability

All relevant data are within the manuscript and its Supporting Information files.

## ACKNOWLEDGEMENTS

The authors would like to thank all participating healthcare workers from ZZG Zorggroep, Zorggroep Maas & Waal and Gasthuis Millingen as well as the healthcare workers who helped with data collection, took the samples and performed the AG-RDTs. We also thank the healthcare workers from the medical microbiology laboratory of Canisius Wilhelmina Hospital for the laboratory work.

